# Towards understanding the disease landscape of clinical trials in Germany: Ontology and embedding-based pipelines versus Large Language Models for ICD-10 Harmonization

**DOI:** 10.64898/2026.08.04.26359616

**Authors:** Rodrigue Ndabashinze, Delwen Franzen, Emilia Kozuch, Jannik Aagerup, Anne Fink, Yerunkar Suresh Samruddhi, Kylie Hunter, Evan Mayo-Wilson, Xiangji Ying, Halil Kilicoglu, Susanne Gabriele Schorr, Anna Lene Seidler

## Abstract

**Background:** Clinical trials conducted in Germany are registered across multiple registries, including the German Clinical Trials Register (DRKS), ClinicalTrials.gov, the EU Clinical Trials Register (EUCTR), and, since 2023, the Clinical Trials Information System (CTIS). These registries record health conditions using different classification systems and terminologies, including ICD-10-GM, MeSH, MedDRA, and free text, making cross-registry analyses difficult. We developed and evaluated a pipeline for harmonizing trial condition descriptions to WHO ICD-10 and compared its performance with that of a large language model (LLM) and to health conditions coded by humans.

**Methods:** We developed a four-stage, registry-aware mapping pipeline consisting of: (i) condition mention extraction and normalization; (ii) classification of ICD-mappable versus non-mappable mentions; (iii) ontology-based candidate generation using UMLS links between MeSH, MedDRA, ICD-10-GM, and WHO ICD-10; and (iv) SapBERT-based semantic retrieval with hybrid confidence scoring. A second variant additionally applied cross-encoder reranking of the top candidate codes. A stratified sample of 500 condition mentions was manually coded to create an expert reference standard. GPT-4o was evaluated in parallel using the same structured decision framework as the human reviewers. Performance was assessed using accuracy, precision, F1 score, and Cohen’s κ at the three-character, block, and chapter levels of ICD-10.

**Results:** The pipeline was applied to 23,061 clinical trials and identified 39,512 ICD-mappable condition mentions, of which 72.4% received a high-confidence assignment. Against 390 expert-coded mentions, the baseline pipeline achieved 49.0% accuracy at the three-character ICD-10 level (κ = 0.487), increasing to 58.7% at the chapter level (κ = 0.561). The cross-encoder method produced small but consistent improvements across all evaluation levels. Candidate-recall analysis showed that the correct code was present in the retrieved candidate set in only 73.7% of cases. The LLM substantially outperformed both pipeline variants, achieving 96.7% accuracy and near-perfect agreement with expert coding (κ = 0.966) at the three-character level. The LLM also assigned clinically plausible codes to 82.4% of rejected mentions, 62.8% of Tier-3 exclusions, and 92.3% of review-band mentions.

**Conclusion:** Automated harmonization of clinical trial condition data across heterogeneous registries is feasible and supports the use of a common ICD-10 framework for cross-registry analyses. The LLMs achieved high agreement with expert coding, and performed better than the deterministic ontology and embedding pipeline, which achieved moderate agreement. These findings indicate that LLMs can support analyses of the distribution of health conditions investigated in clinical trials in Germany.They are a promising tool for classification of other non-standardised trial characteristics in registries.

## 1. Introduction

Identifying health conditions studied in clinical trials is essential for understanding research priorities, identifying gaps in disease coverage, and aligning scientific efforts with national and global health needs^1^. In Germany, clinical trial registration has expanded substantially over the past decade with a gradual increase in prospective registration^2^. While this improves the transparency and reliability of trial registry data, it does not address the inconsistency in how health conditions are captured across trial registries. Registries use different classification terminology. It is therefore difficult to ascertain whether the spectrum of health conditions under investigation in registered trials corresponds to established national and global health priorities, or whether specific disease areas of high public health relevance are underrepresented in clinical trial activity.

Most German clinical trials are registered in the German Clinical Trials Register (DRKS), ClinicalTrials.gov, or the EU Clinical Trials Register (EUCTR). The latter has been transitioning to the Clinical Trials Information System (CTIS) since January 2022 under Regulation (EU) No 536/2014^3^. For the purpose of this study, these two registries will be treated as different registries depending on the year in which the data were collected. These registries use different classification systems for health conditions. DRKS uses the International Classification of Diseases, 10th Revision, German Modification (ICD-10-GM)^4^, whereas EUCTR and CTIS rely on Medical Dictionary for Regulatory Activities (MedDRA) terminology^5^ and ClinicalTrials.gov uses Medical Subject Headings (MeSH) ^6^ terms (for example, ‘Diabetes Mellitus, Type 2’) or free-text condition entries (for example, ‘adult-onset diabetes’). The lack of harmonization complicates cross-registry analysis, limits interoperability, and prevents linkage with national or global disease taxonomies such as the Global Burden of Disease (GBD) classification.

Although manual coding of health conditions into standardized classifications is possible, it is time-consuming, labor-intensive, and subject to inter-coder variability and prone to error^.7^ To address this, previous work explored machine learning approaches to ICD coding and disease classification. Representation learning combining transformers and ontology methods has been shown to achieve high accuracy in mapping medical text to standardized disease categories^8^. Biomedical language models pretrained on PubMed and clinical corpora, including PubMedBERT and SapBERT, have shown promise in clinical concept extraction and entity linking tasks^9–11^. Combining structured ontologies with state-of-the art machine-learning methods can substantially improve how biomedical conditions are identified and matched to standardized concepts^12,13^. For example, embedding-based approaches that generate candidates from established biomedical ontologies and refine them using ranking or similarity-based methods have shown strong performance in biomedical concept normalization and in matching patient- or trial-level information to standardized clinical terminologies. More recently, LLM-based approaches have emerged as a promising extension of these retrieval frameworks^14^. Instead of relying only on lexical matching or embedding similarity, LLMs can leverage broader semantic context from the title of the trial and health condition description, for example to classify that “adult-onset diabetes” and “blood glucose control,” have the same meaning as Type 2 diabetes mellitus.

Our approach applies a two-step mapping strategy. First, we use ontology-based alignment through the Unified Medical Language System (UMLS) to link MeSH and MedDRA terms to WHO ICD-10 codes. Second, for condition texts that cannot be mapped through UMLS, we apply an embedding-based retrieval method that ranks candidate ICD-10 codes according to their semantic similarity to the input text. At the same time, we evaluate an LLM, constrained to the same explicit decision logic used to construct the human reference standard, both as an independent coder for mentions the pipeline cannot confidently resolve as a way to check the pipeline’s high-confidence output.

Performance was evaluated using standard information-retrieval metrics, including precision (the proportion of automated assignments that are correct), recall (the proportion of true conditions that are correctly identified), and the F1 score (the harmonic mean of precision and recall). This work forms an extension to a previous broader project characterizing the landscape of clinical trials conducted in Germany^15^

### Overall aim

To develop and evaluate a reproducible, ontology-driven and embedding-based pipeline for automatically mapping health conditions from German-affiliated clinical trials to WHO ICD-10 classification, using expert manual coding as the reference standard and to compare this performance against a large language model.

### Specific objectives

#### Objective 1

To design and implement a hybrid ICD-10 mapping pipeline that integrates ontology-based candidate mapping via UMLS (MeSH → UMLS → WHO ICD-10, MedDRA → UMLS → WHO ICD-10, ICD-10-GM → WHO ICD-10) with embedding-based retrieval of ICD-10 candidates, and final hierarchical assignment at the three-character category (e.g., E11), with block (e.g., E10–E14) and chapter levels derived programmatically from the assigned code.

#### Objective 2

To evaluate the performance and reliability of the automated pipeline in classifying health conditions into ICD-10 three-character categories, blocks, and chapters, using accuracy, precision, recall, F1 score, and inter-rater agreement metrics, with expert manual coding as the gold standard and to compare this performance against a large language model.

## 2. Methods

### Data sources and study design

We conducted a retrospective, cross-registry harmonization study of clinical trials registered between January 2013 and December 2025 that list Germany as a recruiting country or study site. Data were obtained from the DRKS, ClinicalTrials.gov, the EUCTR, and, from 2023 onward, the CTIS. CTIS and EUCTR predominantly capture trials of regulated medicinal products in the European Union under Regulation (EU) No 536/2014^3^, whereas the DRKS and ClinicalTrials.gov include a broader range of interventional studies; accordingly, the combined registry was not restricted to regulated products alone. The study included all registration versions available at the time of data extraction, not limited to prospective registrations. Registry fields used for health condition extraction were restricted to condition-designated fields (condition names, ICD codes, MeSH terms, therapeutic areas); eligibility criteria, outcome measures, and other fields were not used as primary disease sources to reduce the risk of capturing conditions that are exclusion criteria or endpoints.

### Cross-registration linkage and harmonization

Trials registered in more than one registry were identified using a hierarchical linkage method: exact registry and secondary identifiers first (DRKS IDs, NCT numbers, EudraCT/CTIS identifiers), then normalized exact title matching, then approximate title matching when identifiers were absent or inconsistent, in line with previously validated cross-registration linkage methods^16^.

For linked trials, health condition fields used for ICD-10 mapping followed a distinct, field-level rule: the structured code field was populated from DRKS’s ICD-10-GM code where present, falling back to ClinicalTrials.gov MeSH code when DRKS code was absent (CTIS and EUCTR did not contribute to this specific field). CTIS’s own MedDRA condition terms were used directly via the CTIS/EUCTR pathway. We used free-text condition descriptions from each registry condition field, falling back to CTIS’s Medical Conditions field only when that field was empty; and CTIS’s therapeutic-area field, where available, was retained separately as chapter-level context (Pathway D). Condition mentions from all linked records were merged and deduplicated.

### Data preprocessing and multi-field integration

Information about health conditions is captured in different formats across registries. DRKS provides structured free-text condition descriptions alongside ICD-10-GM codes. ClinicalTrials.gov uses a mixture of MeSH-coded conditions and free-text descriptions. EUCTR and CTIS record medical conditions using MedDRA terminology and therapeutic area metadata, usually at the Preferred Term (PT) level, often containing investigator-entered free text. In this study, MedDRA is used as an interoperability vocabulary for condition mapping rather than as an adverse-event report; the CTIS and EUCTR condition fields are distinct from results-reporting adverse event fields.

The pipeline uses registry field names to distinguish condition-related text from information about interventions, outcomes, or eligibility instead of treating all fields as equivalent sources of disease information. Condition-specific fields serve as the primary inputs for disease extraction. Official titles and, when available, brief summaries are used only as contextual support for disambiguation. Intervention, endpoint, results, and administrative fields are excluded as primary disease sources to avoid contamination from exclusions, procedures, healthy-volunteer labels, adverse events, or other content that does not represent the trial’s target health condition.

Condition strings containing more than one health problem (for example, “type 2 diabetes; obesity”, or “Crohn’s disease and ulcerative colitis”) were split into individual mentions using high-precision delimiter rules where splitting remains semantically defensible. Each resulting mention was processed independently and later re-aggregated at the trial level so that multi-label ICD-10 profiles can be retained.

Text normalization preserved the original mention string while generating a normalized mention representation used for matching and retrieval. Normalization included lowercasing, whitespace normalization, limited punctuation cleanup, harmonization of common clinical abbreviations and acronyms, German-to-English translation of frequently occurring disease terms and synonym consolidation.

### Overview of the reproducible four-stage mapping pipeline

The primary analytical unit is the individual condition mention. The pipeline is registry-aware and attempts deterministic ontology mappings first, with embedding-based retrieval contributing to every mention’s hybrid confidence score and serving as the sole basis for assignment only when no ontology-derived candidate exists.

Figure 1 below summarizes the workflow from mention extraction and normalization (Stage 1), through mention-level ICD eligibility classification (Stage 2) and registry-specific ontology mapping (Stage 3), to embedding-supported ranking, hierarchical ICD-10 assignment, and trial-level aggregation (Stage 4). Each stage is described in detail below, in the order shown

**Figure 1.**
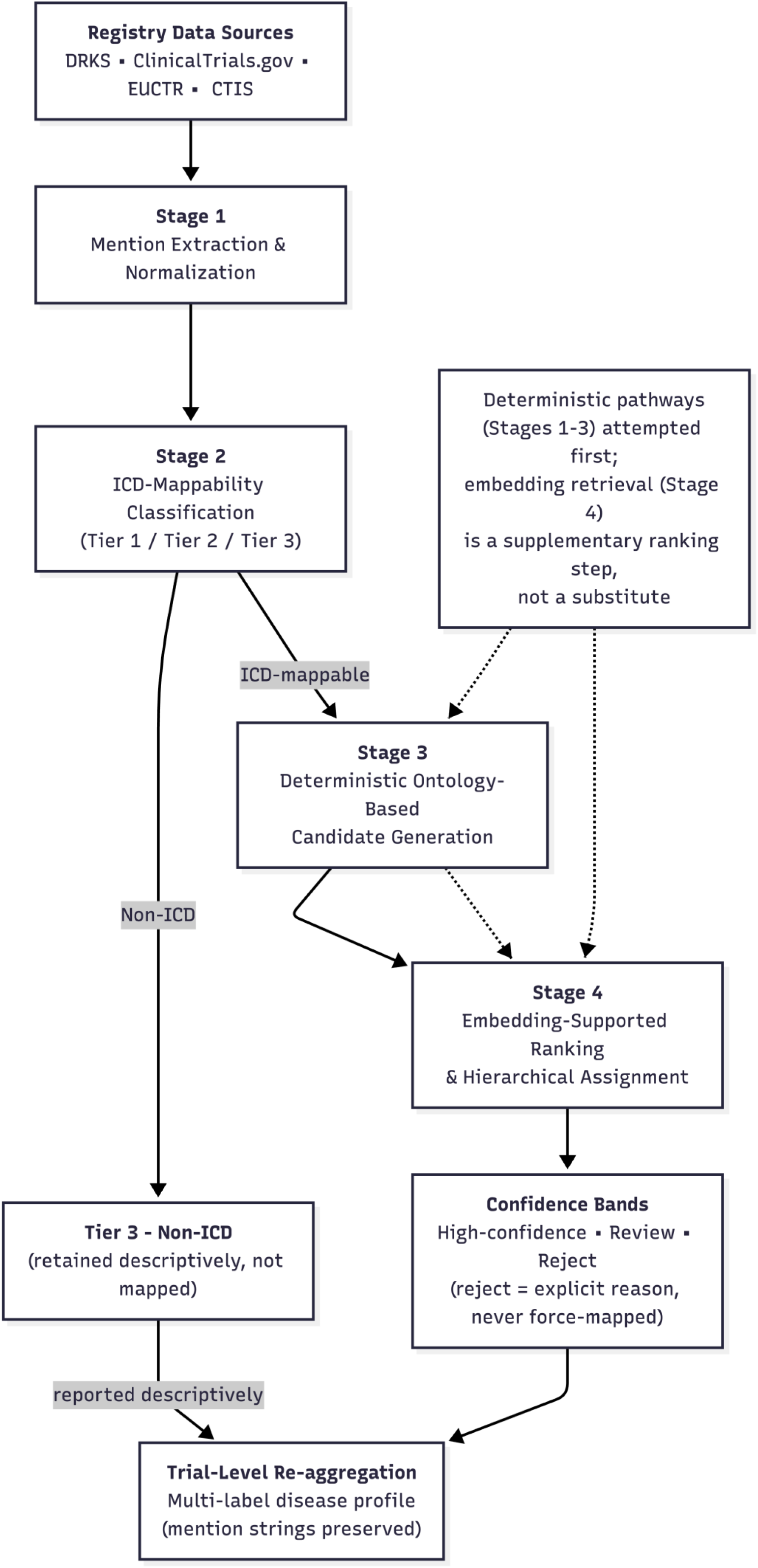
Overview of the four-stage ICD-10 mapping pipeline. Condition mentions extracted from four clinical trial registries are processed through mention extraction and normalization (Stage 1), ICD-mappability classification (Stage 2), deterministic ontology-based candidate generation (Stage 3), and embedding-supported ranking with hierarchical assignment (Stage 4), before aggregation into trial-level multi-label disease profiles.

### Stage 1: Mention extraction and normalization

Each condition field is parsed into individual mentions, its registry source and stored with raw text, normalized text, harmonized trial identifier, and any structured terminology already present in the record (e.g., a DRKS ICD-10-GM code). A separate rescue step identifies condition fields that contain narrative study descriptions rather than actual condition terms. Using a heuristic rule, fields exceeding 200 characters and containing study-description keywords (e.g., “trial”, “evaluate”, or “efficacy”) are classified as narrative text. When such records have an associated structured condition code, that code is substituted for the narrative text and carried forward for subsequent mapping.

For DRKS entries, where a mention has a structured ICD-10-GM code, it is carried forward alongside the mention text as paired evidence for embedding-based retrieval.

### Stage 2: Mention-level ICD classification

Stage 2 distinguishes mentions that are eligible for ICD-10 mapping from non-ICD concepts. ICD-mappable mentions include Tier 1 disease entities (codes in chapters A–Q and U-COVID) and analytically relevant Tier 2 ICD concepts when they are the explicit condition under study, such as symptoms (R-codes), injuries (S/T-codes), pregnancy-related conditions (O-codes), and encounters or status codes (Z-codes). Non-ICD concepts (Tier 3) are mentions that do not correspond to any ICD-10 code; for example trials in healthy volunteers, pharmacokinetic or bioequivalence studies, methodological or basic-research trials.

Classification followed a priority hierarchy: a global stop-list first removed text that is clearly not a health condition regardless of context (bare anatomical terms, drug-exposure descriptions with no disease signal); structured ICD-10-GM disease-chapter codes are then labelled deterministically ICD-mappable; structured Tier 2 codes (R/S/T/O/Z) undergo a contextual check against trial title/context, since the presence of such a code does not guarantee it represents the condition under study; explicit Tier 3 phrase patterns are labelled deterministically non-ICD. Mentions with no structured code guidance at all were passed to a supervised elastic-net logistic regression classifier.

The classifier was trained on silver standard labels drawn from the deterministically-labelled mentions, using TF-IDF unigram/character n-gram features, mention length, registry indicators, and primary purpose category. Because disease mentions are more frequent than non-disease mentions, the classifier uses inverse-frequency class weighting, and its decision threshold was calibrated on a held-out set to maximize F1 for the minority Tier 3 class. A conservative disease-keyword override deterministically reclassifies any mention containing an unambiguous disease term (e.g., cancer, diabetes, stroke, depression, asthma, obesity) that the classifier had labelled Tier 3. The formal elastic-net specification and decision rule are provided in Appendix A2.

### Stage 3: Deterministic ontology-based mapping via UMLS

For mentions classified as ICD-mappable, Stage 3 generates candidate WHO ICD-10 three-character codes via deterministic terminology pathways.

**DRKS pathway:** the ICD-10-GM code is carried forward, extracting the shared three character code between ICD-10-GM and ICD-10-WHO (the two systems are identical at this level for the majority of categories by design, since ICD-10-GM is a German extension of ICD-10-WHO).

**ClinicalTrials.gov pathway:** structured MeSH terms and free-text condition strings linked through UMLS Concept Unique Identifiers (CUIs) to ICD-10 with parent and ancestor terms extracted from the MeSH hierarchy also considered as additional mapping candidates.

**CTIS/EUCTR pathway:** MedDRA condition strings normalized and linked through UMLS, with the CTIS therapeutic-area field used only as a weak chapter-level prior for consistency check; and a string fallback pathway using Jaro-Winkler approximate matching (similarity threshold δ = 0.80), used only when no candidates are generated by other ontology pathways.

We used UMLS semantic types (from MRSTY)17to filter concepts that are not disease terms and gave high priority to candidates that look like broad diseases (cancers, mental-disorder, congenital etc); candidates in the R/Z code range are excluded from this scoring pool entirely, since Tier 2 R/Z codes are resolved through the separate deterministic Stage 2 pathway. For DRKS records containing structured ICD-10-GM codes, a specific code was retained directly as supporting evidence. When the associated code represents a range or chapter-level expression rather than a single category (e.g., C00-C14), the range was expanded into all valid WHO ICD-10 codes in that range and retained as broad, lower-confidence evidence, provided that the expansion yielded no more than 25 codes. Wider ranges, including common chapter-level expressions such as C00-C97, were not expanded. Instead, the first three characters of the range expression were used as a fallback candidate. In practice, this means a broad, non-specific range entered by the trial registrant may contribute evidence that reflects only the first code in the reported range, rather than the underlying condition described by the mention.

(Appendix A.3 provides the formal candidate set definition and the confidence scores assigned to each pathway)

### Stage 4: Embedding-supported ranking and hierarchical assignment

SapBERT embeddings were computed for each mention and for a curated ICD-10 concept index in which every three-character WHO code is represented by its official English label, UMLS linked MeSH/MedDRA synonyms, and its block/chapter labels. FAISS approximate nearest-neighbour search^18^ (a library for efficient similarity search in high-dimensional vector spaces) retrieved the top-10 semantically similar candidates per mention.

Deterministic (Stage 3) and embedding (Stage 4) evidence are combined into a hybrid confidence score. Two distinct formulas are used, depending on whether a candidate has ontology support.

For candidates with ontology support:

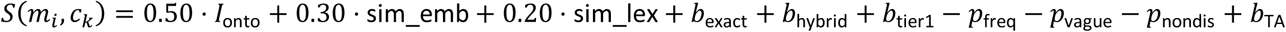

For candidates with no ontology support at all (embedding-only mentions), a separate, simpler formula applies instead:

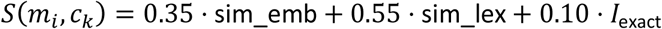

Where :

*I*_onto_: The Stage 3 ontology confidence score (zero for embedding-only candidates).

sim_emb: The SapBERT cosine similarity between the mention and the candidate embedding.

sim_lex: The Jaro-Winkler lexical similarity measuring string-level match.

*I*_exact_: An indicator (0 or 1) for an exact string match, used directly in the embedding-only formula.

*b*_exact_: A bonus of 0.15 (ontology-backed formula only) applied if there is an exact string match between the mention and the concept label.

*b*_hybrid_: A bonus of 0.03 applied when both ontology and embedding evidence support the same candidate.

*b*_tier1_: A bonus of 0.05 applied for Tier 1 disease-chapter candidates.

*p*_freq_: A penalty proportional to the log-frequency of how often a code has already been assigned elsewhere in the corpus, reducing over-reliance on common attractor codes.

*p*_vague_: A fixed penalty of 0.15 applied to a curated blocklist of known vague “attractor” codes (C80, R69, Z00, D48, T98) that otherwise attract unrelated mentions in high-dimensional embedding space.

*p*_nondis_: A penalty of 0.15 applied when the mention text matches a non-disease textual pattern.

*b*_TA_: For CTIS mentions specifically, a bonus of 0.08 or penalty of 0.10 depending on whether the candidate chapter agrees with the trial’s corresponding therapeutic area.

The winning candidate is defined as :

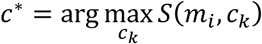

For each mention, a hybrid confidence score, *S*(*m_i_*, *c_k_*), is computed for every candidate ICD-10 code, where *m_i_* denotes the mention being mapped and *c_k_* denotes a candidate code. The final assignment is determined as *c*^∗^ = arg max *S* (*m_i_*, *c_k_*), where the pipeline selects the candidate code that achieves the highest confidence score among all candidates. *c*^∗^ represents the code judged to be the best overall match for the mention based on all available evidence. Where the top two candidates fall within the same ICD-10 block and their scores differ by less than 0.03, the pipeline assigns a pre-specified family-level default code (e.g., unspecified diabetes for an ambiguous diabetes mention or unspecified malignant neoplasm for an ambiguous cancer mention) instead of inferring a specific subtype. This choice maximizes precision over specificity when the available evidence does not clearly distinguish between related codes.

A separate curated list of known disease-name patterns can override the selected candidate for a small number of mentions. For example, if a mention matches a predefined pattern such as “COVID-19”, the pipeline directly assigns the corresponding ICD-10 code from the curated mapping without using the highest-scoring candidate. In such cases, the assigned code, label, and confidence score are replaced by the predefined values, and the confidence score is fixed at 0.85.

Each mention is then assigned to one of three confidence bands. High-confidence assignments require either an ontology-supported final score of at least 0.50 or, for embedding-only candidates, the predefined thresholds for cosine similarity, lexical similarity, and final score. A review band captures borderline ontology-supported assignments and embedding-only candidates. All remaining mentions are assigned to the reject band, accompanied by an explicit reason (e.g., no candidate identified, low embedding signal, insufficient evidence, or range-code-only evidence).

An alternative Stage 4 architecture (“Method 2”) applied a BAAI/bge-reranker-base cross-encoder to rerank the top-10 SapBERT candidates before hybrid scoring; the reranked score is trusted in place of cosine similarity only when the cross-encoder confidence exceeds a fixed threshold, and reverts to the original SapBERT ranking otherwise.

Method 2 is compared against baseline throughout the Results.

Once a three-character code is retained, its ICD-10 block and chapter are derived programmatically from the official WHO hierarchy table.

**Figure 2** illustrates a worked example of this hierarchical assignment process on a single trial

**Figure 2.**
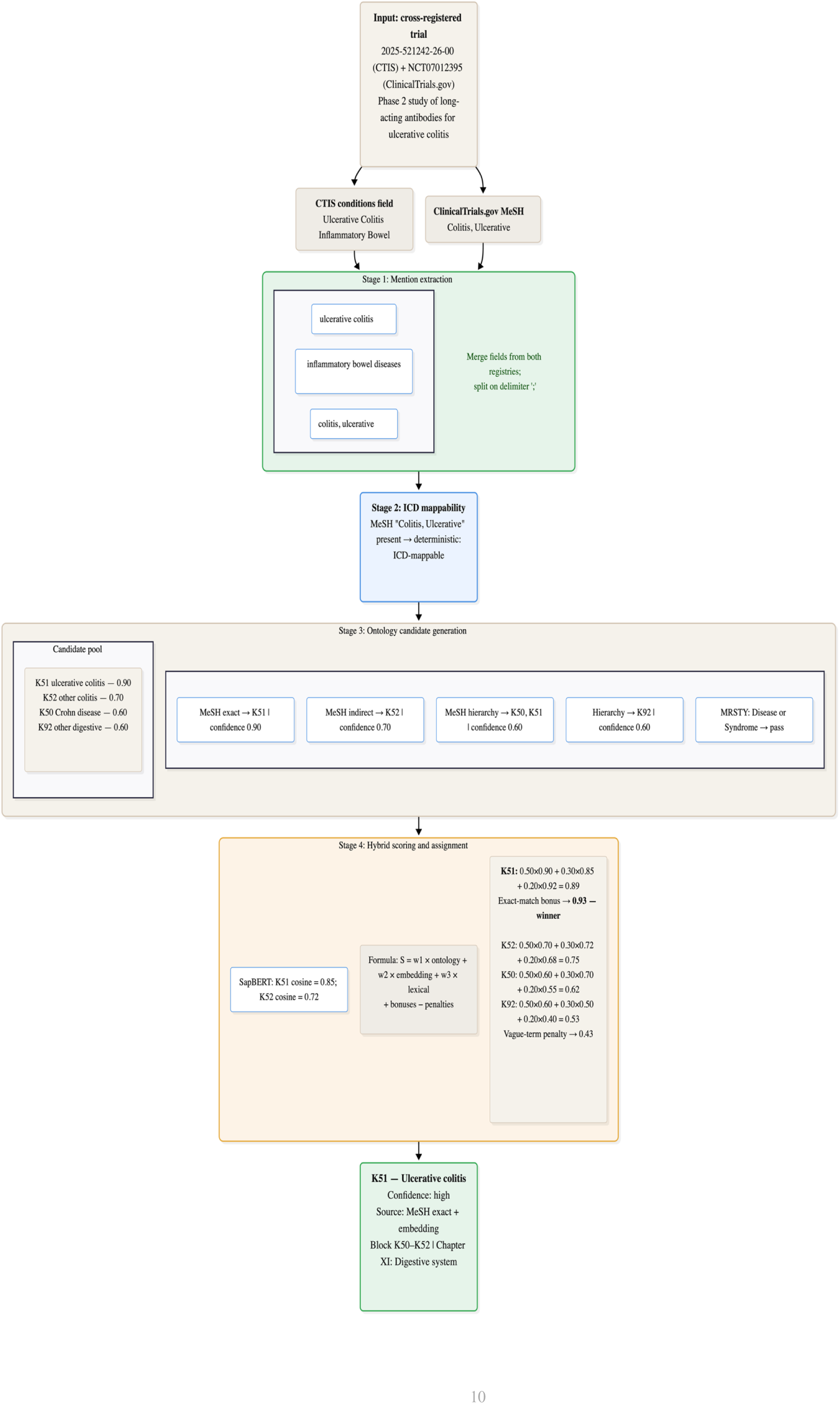
Worked example of the four-stage mapping pipeline on a cross-registered trial, illustrating how condition fields are merged, split into mentions, classified, matched to candidate codes, and scored to select the final high-confidence assignment

### Trial-level aggregation

Mention-level output is the primary analytical unit, but trial-level summaries are derived by aggregating all retained mention-level codes within a trial into a multi-label set (high-confidence Tier 1 codes, high-confidence Tier 2 codes, review-band codes, and retained Tier 3 labels where applicable). Trial-level agreement, where reported, is evaluated using Jaccard similarity between predicted and true code sets. Mentions assigned to the review or reject bands are retained as explicit outputs. Review-band mentions are exported for structured manual checking, while rejected mentions are exported with deterministic reject reasons and suggested actions.

### LLM comparison

#### Track A (expert gold standard)

We selected a stratified random sample of 500 condition mentions from the review band, reject band, embedding-only assignments, and Tier 3 mentions. Two medical reviewers independently coded a 50-mention calibration subset at the three-character ICD-10 level. A pre-specified inter-rater agreement threshold of κ ≥ 0.70 was used to assess consistency between reviewers. If this threshold was achieved, the remaining mentions were independently coded; otherwise, all mentions underwent dual review. GPT-4o (OpenAI; temperature = 0; structured JSON output) was used as a supplementary adjudicator. The model was applied to all review-band mentions, reject-band mentions, sub-threshold embedding-only assignments, and Tier 3 mentions. To align annotator and LLM-based assessment, GPT-4o followed the same decision cascade and annotation framework used by the medical reviewers. Specifically, both human reviewers and the LLM first attempted direct ICD-10 assignment from the mention text, then considered additional trial context where necessary, and finally applied broader GBD-based classifications when an exact ICD-10 mapping could not be identified. The annotation guide provided to human reviewers is described in Appendix B.5; Appendix B.1.

#### Track B - LLM-based precision check of the high-confidence band

To provide an additional assessment of pipeline accuracy, we drew a stratified sample of 250 mentions from the high-confidence band, with sampling distributed across ICD-10 chapters. Each mention was independently reviewed by GPT-4o, which returned one of four structured outcomes: confirm, correct, flag, or no_code. To align manual and LLM-based evaluation, GPT-4o and human reviewers followed the same decision framework. First, reviewers assigned an ICD-10 code directly from the condition mention whenever the mention was sufficiently specific. If the mention alone was insufficient, the reviewer considered additional trial information, including the study title and registration record, to infer the most appropriate condition. When no exact ICD-10 code could be assigned, reviewers mapped the condition to the corresponding GBD Level 3 category. If no suitable Level 3 code existed, the Level 3 category name was recorded without a code. When a Level 3 classification was not possible, the reviewer assigned the broader GBD Level 2 category. A NO_ICD decision was used only when all preceding steps failed to produce a valid classification.

Two predefined rules were applied to ensure consistent handling of ambiguous cases. For prevention studies and healthy-population cohorts, coding was based on the disease or condition being prevented rather than the healthy population under investigation. In addition, when multiple coding options were available, preference was given to the underlying disease over isolated symptoms and to the primary cause of a condition over its clinical manifestations.

### Evaluation and performance

The primary unit of evaluation was the individual condition mention, assessed at the exact three-character ICD-10 level. Performance at the ICD-10 block and chapter levels was derived deterministically from the retained three-character code. At these higher levels, predictions were considered correct when the predicted and reference codes belonged to the same block or chapter, even if the exact three-character codes differed. This approach acknowledges that errors between clinically related codes within the same hierarchy are less severe than errors with different ICD-10 chapters.

Performance was evaluated using recall (sensitivity), precision, F1-score, and unweighted Cohen’s κ. Recall was defined as *TP*/(*TP* + *FN* + *Wrong*), precision as *TP*/(*TP* + *Wrong* + *FP*), and the F1-score as the harmonic mean of precision and recall. Cohen’s κ was calculated as *κ* = (*p_o_* − *p_e_*)/(1 − *p_e_*), where *p_o_*represents the observed agreement between automated and reference classifications and *p_e_*represents the agreement expected by chance based on the marginal category distributions.

Cohen’s κ was computed using the scikit-learn implementation (cohen_kappa_score) and interpreted according to the Landis and Koch framework^19^: slight (0.00 - 0.20), fair (0.21-0.40), moderate (0.41-0.60), substantial (0.61-0.80), and almost perfect agreement (0.81-1.00).

Performance was evaluated separately across confidence bands (high-confidence, review, and reject) and across mapping strata (ontology-supported, embedding-only, Tier 3, and unspecified-default assignments) to identify areas of strong performance and those requiring additional review. As a secondary evaluation, candidate recall was assessed independently of the final ranking step. Specifically, we measured whether the expert-assigned ICD-10 code appeared anywhere within the retrieved candidate set. This analysis was conducted separately for ontology-derived and embedding-derived candidates to differentiate errors arising during candidate generation from those arising during candidate ranking.

The SapBERT method and Method 2 were compared descriptively at each ICD-10 hierarchy level using the evaluation metrics described above. In addition, a row-level agreement analysis was performed by comparing the three-character ICD-10 predictions generated by the two methods for the same mentions. This analysis was intended to determine whether observed performance differences reflected different coding decisions or differences in abstention behavior.

The LLM-based evaluation tracks (Track A and Track B) were assessed against the same expert reference standard using the same evaluation framework for direct comparison between the deterministic mapping pipeline and the LLM-constrained decision process across all ICD-10 hierarchy levels.

### Error analysis

Disagreements between the automated pipeline and the expert reference were classified using a pre-defined error taxonomy. Error categories included false positives, false negatives, ontology-link failures (where no valid UMLS-based mapping could be established), embedding attractor errors (where semantically broad or highly connected codes attracted unrelated mentions), within-family ambiguities (incorrect assignment of a subtype within the correct ICD-10 block), cross-chapter misclassifications (assignment to an incorrect disease domain), and retention of non-condition text treated as a condition mention.

To distinguish within-family from cross-chapter errors, we used the official ICD-10 chapter structure rather than comparing only the three-character codes. This was important because some ICD-10 chapters cover more than one code range. For example, the Neoplasms chapter includes both C00-C97 and D00-D48. As a result, a mismatch between a C-code and a D-code may still occur within the same ICD-10 chapter. Using the official ICD-10 hierarchy ensured that related coding errors were classified correctly and were not counted as cross-chapter errors.

## 3. Results

In total, we processed 23,061 German-affiliated interventional trials, from which 42,838 individual condition mentions were extracted during Stage 1. Among these, 226 mentions (0.5%) that did not describe a health condition in any context were excluded, leaving 42,612 eligible condition mentions for analysis. The following result sections present the results obtained at each stage of the pipeline. First, we describe the outcomes of the condition extraction process in Stage 1. Second, we report the performance of the classifier used in Stage 2. Finally, we present and compare the performance of the baseline SapBERT bi-encoder model, the cross-encoder reranker (Method 2), and the LLM against the gold-standard annotations.

### 3.1 Stage 1: Condition-mention extraction

Of the 42,612 eligible mentions, deterministic rules resolved 33,713 (79.1%) without model involvement; the remaining 8,899 (20.9%), lacking structured guidance, were passed to the Stage 2 classifier.

**Table 1.** Stage 1 condition-mention routing prior to classifier (N = 42,612 eligible mentions).

| Pathway | N (mentions) | ICD-mappable | Tier 3 |
| --- | --- | --- | --- |
| Structured Tier 1 (chapters A–Q, U-COVID) | 32,584 | 32,584 | 0 |
| Structured Tier 2 (contextually checked) | 1,125 | 545 | 580 |
| Explicit Tier 3 pattern | 4 | 0 | 4 |
| No structured guidance (Stage 2 classifier) | 8,899 | 6,383 | 2,516 |
| Total | 42,612 | 39,512 | 3,100 |

The following section reports performance on a held-out test split and then describes how classifier outputs were combined with the deterministic pathways to generate the ICD-mappable mentions passed to Stage 3.

### 3.2 Stage 2: Classifier performance

**Table 2.** Elastic net classifier performance.

| Metric | Value |
| --- | --- |
| Overall accuracy | 91.3% |
| Recall ICD-mappable class | 93.5% |
| Recall Tier 3 (minority) class | 31.9% |
| AUC | 0.783 |

The high overall accuracy and ICD-mappable recall indicate that the classifier reliably identifies disease mentions. Tier-3 recall is lower; the classifier misses two in three non-disease mentions in the held-out test set, calling them ICD-mappable instead. The decision threshold was intentionally calibrated to maximize the F1 score of the minority Tier 3 class, with an emphasis on preserving true disease mentions.

### 3.3 Stage 3 – 4: Ontology and embedding evidence for the overall corpus

Across the 39,512 ICD-mappable condition mentions, ontology evidence supported 87.7% of final assignments through either ontology-only or hybrid pathways. Embedding retrieval was the sole source of evidence for 11.3% of assignments. SapBERT and the cross-encoder produced similar proportions of high-confidence assignments (72.4% and 71.8%, respectively) because confidence classification was based primarily on ontology evidence. The cross-encoder reranked candidates generated by the pipeline but did not change the evidence available for confidence classification. Consequently, its effect is not reflected in coverage statistics and is assessed through comparison with Tables S1–S3.

### 3.4 Performance against Gold-standard

For the 50-mention calibration subset, the inter-rater agreement pre-specified threshold of κ ≥ 0.70 was achieved (κ = 0.751 for three-character ICD-10 codes and κ = 0.821 for ICD-10 chapters). The reviewers then independently annotated the remaining 450 condition mentions. Table 3 summarizes the composition of the evaluation corpus.

**Table 3.** Composition of the evaluation corpus (N = 500 condition mentions)

| Design stratum | n | % of total | Expert-coded |
| --- | --- | --- | --- |
| Ontology-supported | 200 | 40% | 197 |
| Embedding-only | 150 | 30% | 93 |
| Tier-3 excluded | 100 | 20% | 51 |
| Unspecified default | 50 | 10% | 49 |
| Total | 500 | 100% | 390 |

Of the 500 sampled condition mentions, experts confirmed that 390 (78.0%) represented codable health conditions. Confirmation rates differed across strata, ranging from 98.5% (197/200) for ontology-supported mentions down to 51.0% (51/100) for Tier-3-excluded mentions, indicating that the pipeline’s confidence levels corresponded closely with expert assessment. The Tier-3-excluded stratum therefore retained a substantial minority of codable conditions despite being excluded by the pipeline. All subsequent accuracy analyses were restricted to the 390 expert-confirmed condition mentions.

### 3.5 Three-way comparison against the expert reference standard

Table 4 compares agreement with the expert reference standard across ICD-10 hierarchy levels. The baseline and Method 2 achieved similar performance, with modest improvements at broader hierarchy levels. In contrast, the LLM achieved consistently high accuracy and agreement across all levels, reaching 96.7% accuracy and κ = 0.966 at the three-character level. Detailed confusion-matrix results are provided in Supplementary Table S4.

**Table 4.**
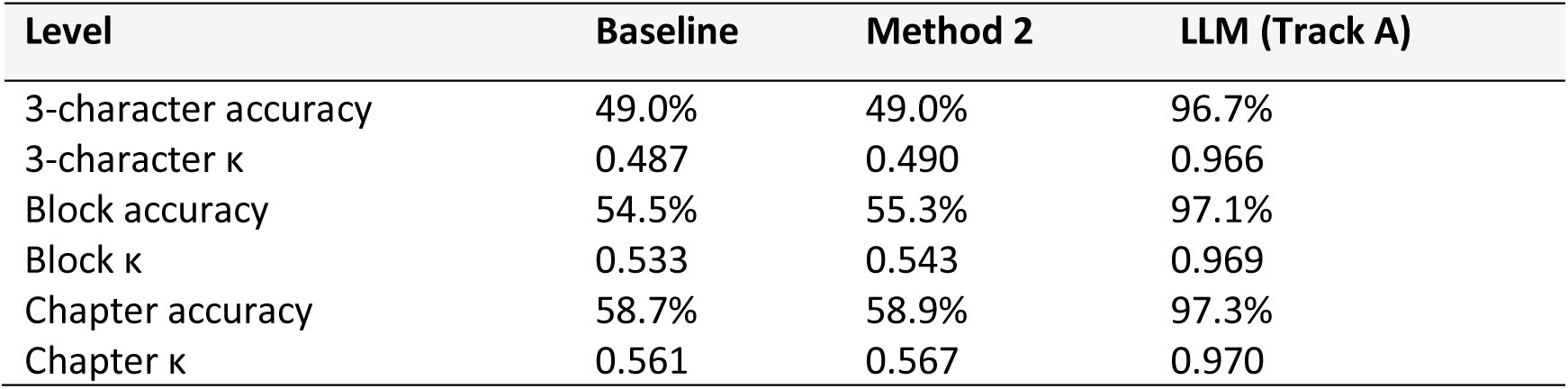
Accuracy and Cohen’s κ against the expert reference standard, by method and hierarchy level (LLM figures reflect adjudicated results)

| Level | Baseline | Method 2 | LLM (Track A) |
| --- | --- | --- | --- |
| 3-character accuracy | 49.0% | 49.0% | 96.7% |
| 3-character $\kappa$ | 0.487 | 0.490 | 0.966 |
| Block accuracy | 54.5% | 55.3% | 97.1% |
| Block $\kappa$ | 0.533 | 0.543 | 0.969 |
| Chapter accuracy | 58.7% | 58.9% | 97.3% |
| Chapter $\kappa$ | 0.561 | 0.567 | 0.970 |

**Figure 3.**
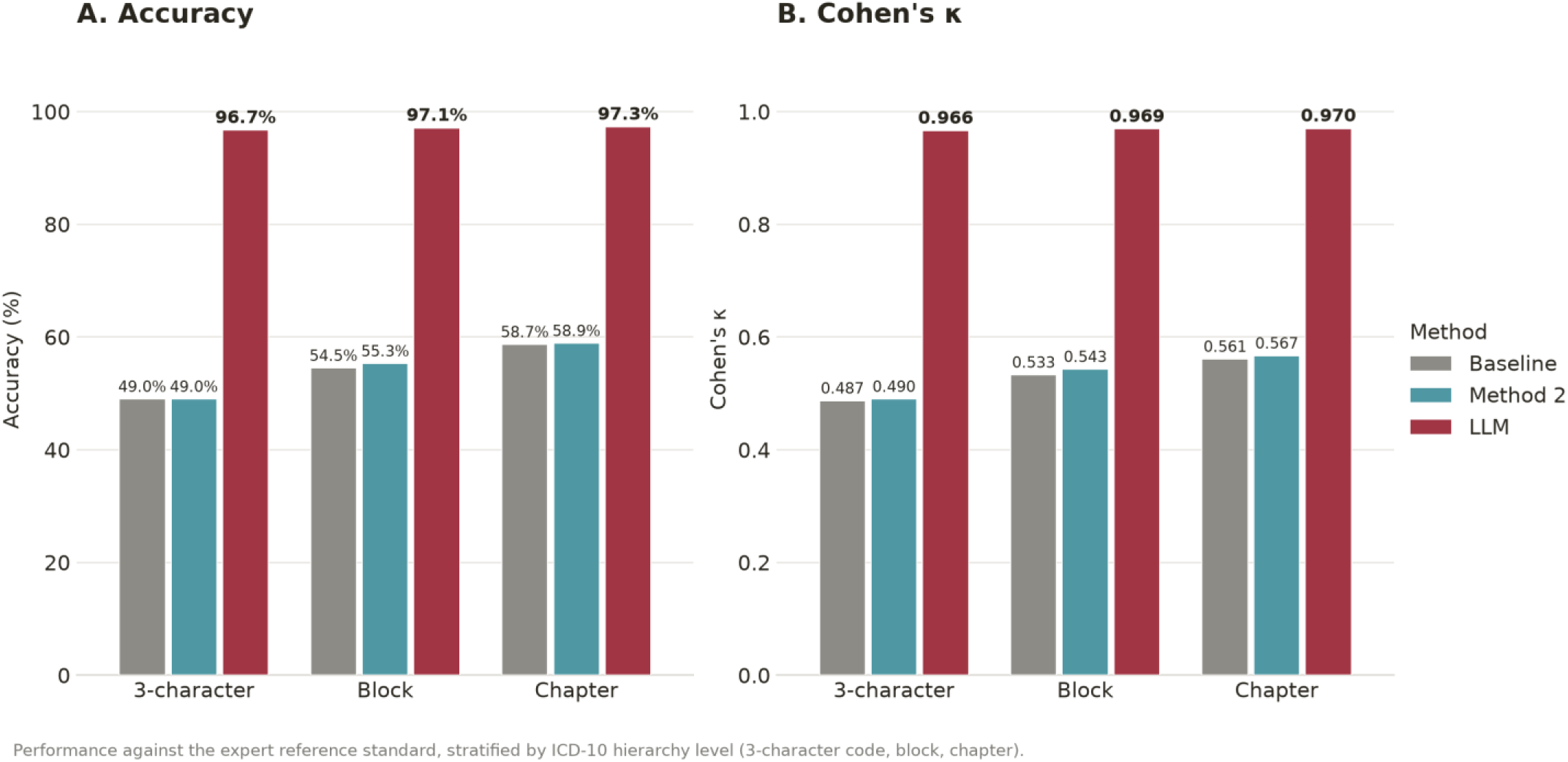
Accuracy and Cohen’s κ against the expert reference standard, by method and hierarchy level. LLM figures reflect adjudicated results

Panel A shows accuracy and Panel B shows Cohen’s κ for the baseline method, Method 2, and the LLM. The LLM achieved substantially higher agreement with the expert reference standard across all hierarchy levels.

### 3.6 LLM performance: adjudication and recovery of pipeline failures

Table 5 shows that adjudication substantially improved agreement between the LLM and the expert reference standard. After expert review of disagreements, three-character accuracy increased from 62.7% to 96.7%, and Cohen’s κ increased from 0.625 to 0.966. When applied to cases not resolved by the deterministic pipeline, the LLM assigned ICD-10 codes to 82.4% of reject-band mentions, 62.8% of Tier 3 mentions, and 92.3% of review-band mentions.

**Table 5.**
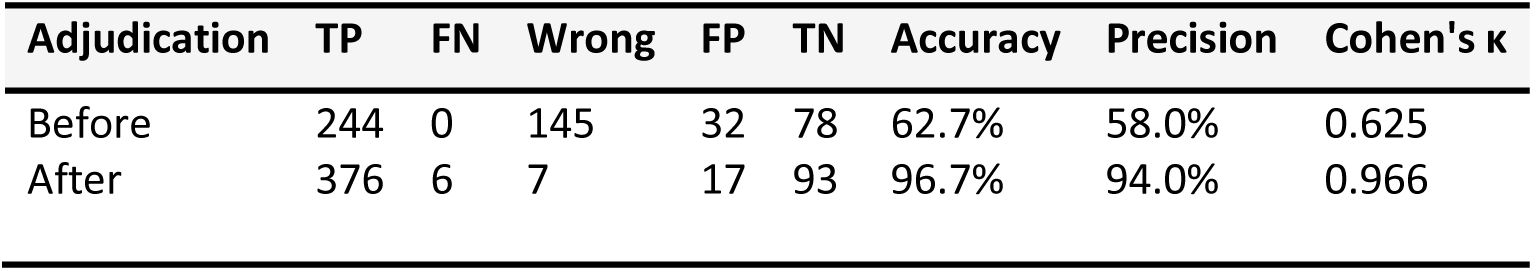
LLM (Track A) confusion matrix, raw vs. adjudicated (3-character level).

**Figure 4.**
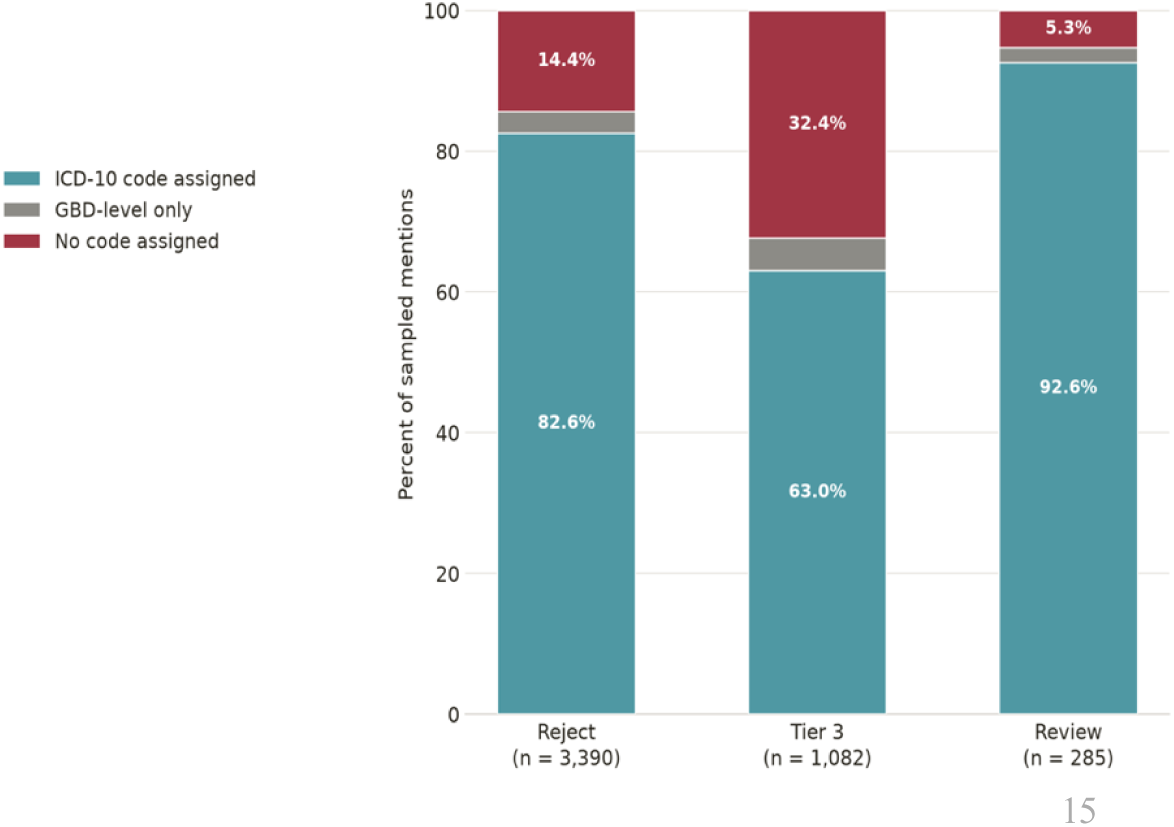
LLM recovery of condition mentions not resolved by the deterministic pipeline. Stacked bars show the proportion of mentions for which the LLM assigned an ICD-10 code, assigned only a GBD-level category, or agreed that no code was warranted across the reject, Tier 3, and review bands.

### 3.7 Error analysis of the pipeline

Most errors were found to occur between related conditions within the same disease family or chapter (64.3% for the baseline method and 66.1% for method 2. In contrast, true cross-chapter errors, where the predicted code belonged to a different disease domain, accounted for 30.4% and 28.6% of errors, respectively. This distinction was important because a simple code-prefix approach can misclassify related errors as cross-chapter disagreements. For example, the ICD-10 neoplasm chapter includes both C-codes (C00-C97) and D-codes (D00-D48), meaning that some C-code-to-D-code transitions are incorrectly classified as cross-chapter errors under a naive first-letter comparison, when they are in fact within-chapter.

Findings also indicated that approximately one-quarter of coding errors came from candidate generation, where the correct code was never retrieved and therefore could not be recovered during reranking. Performance depended mostly on the availability of ontology evidence. When an ontology-based mapping was available, accuracy reached 79.2%; for embedding-only mentions, accuracy fell to 1.1%. Detailed error analyses, candidate-recall results, and reliability stratifications are provided in Supplementary Note 1 and Supplementary Tables S5–S6.

## 4. Discussion

As part of a larger effort to characterize the landscape of clinical trials conducted in Germany, we developed and evaluated an automated pipeline for harmonizing health conditions reported across four major trial registries (ClinicalTrials.gov, DRKS, EUCTR, and CTIS) to a common WHO ICD-10 classification. The proposed framework combines deterministic ontology-based mappings through UMLS with embedding-based semantic retrieval. We also evaluated the performance of a LLM against the pipeline using the same decision logic used to construct the human reference standard.

The findings indicate that the LLM, evaluated in parallel against the deterministic pipeline, achieved substantially higher performance when constrained to the same decision logic with the human reference standard. The LLM achieved an adjudicated accuracy of 96.7% and near-perfect agreement with expert review (κ = 0.966) at the exact three-character ICD-10 level. This exceeded the performance of the deterministic pipeline, which achieved 49.0% accuracy, with moderate agreement with expert coding (κ = 0.487).

The advantage of the LLM was not limited to broad disease categories. Its performance remained almost unchanged across the three-character, block, and chapter levels of ICD-10, indicating that it was correctly resolving errors at the most specific coding level. The LLM also successfully assigned clinically plausible codes to many mentions that the deterministic pipeline could not classify with confidence, including 82.4% of reject-band mentions, 62.8% of Tier-3 exclusions, and 92.3% of review-band mentions. These findings suggest that many failures of the deterministic pipeline came from limitations in terminology matching and candidate retrieval.

These results are consistent with growing evidence that LLMs are most effective when used within structured coding frameworks. Akkhawatthanakun et al.^20^ found that LLMs could not perform well when asked to generate complete ICD code sets independently, but achieved much better results when reviewing and refining candidate codes generated by a supervised deep-learning coder. Similarly, Vassileva et al.^21^ showed that combining GPT-4.1 with dictionary-based candidate retrieval outperformed their own standalone LLM baselines for multilingual ICD-10 entity linking. These studies suggest that ontology and retrieval methods are important for generating candidate codes, while LLMs contribute most effectively by reviewing, clarifying, and selecting among clinically plausible alternatives.

Anjos de Almeida et al.^22^ reached a similar conclusion in a large benchmark of medical code selection systems. 10 of the 33 evaluated models achieved F1 scores above 0.85, and performance improved further when the correct code was guaranteed to be present in the candidate set. This finding highlights the importance of candidate retrieval quality: even highly capable models cannot select the correct code if it was never retrieved in the first place. Increasing evidence therefore suggests that the greatest value of modern LLMs lies in clinical reasoning, disambiguation, and candidate evaluation within a constrained set of plausible diagnoses instead of generating ICD codes from a prompt without external support.

Although retrieval-based methods generally improve coding performance, the size of the improvement varies depending on the task and model used. For example, Gan et al.^23^ reported a 3-13% improvement in micro-F1 after incorporating structured external knowledge into an LLM-based coding system. Their approach achieved performance comparable to state-of-the-art supervised methods, demonstrating that external knowledge can provide substantial benefits under the right conditions.

Differences across studies are likely explained by differences in task complexity. Retrieval methods tend to provide the greatest benefit when condition descriptions are clear and the number of possible codes is small. In contrast, improvements are often smaller in complex coding tasks involving long clinical narratives and thousands of possible diagnoses. This interpretation is consistent with the findings of the present study. Although Method 2 produced only small improvements in overall accuracy and precision, these gains should be viewed in the context of the candidate-recall ceiling. Re-ranking methods can only choose among codes that have already been retrieved. If the correct code is absent from the candidate set, no ranking method can recover it. Consequently, a more informative measure of improvement was the reduction in unspecified default classifications, with D48 assignments decreasing from 29 to 11 among the same set of 113 mentions. This suggests that the method became better at distinguishing between closely related diagnoses once appropriate candidates had been retrieved. In our study, the performance of the LLM further supports this interpretation. The model was not asked to generate ICD codes freely from its own knowledge. Instead, it operated within a structured decision framework based on the same rules used to create the human reference standard. This result places the present study among the highest-performing examples of grounded LLM-assisted coding systems and highlights the continued importance of external knowledge and structured prompts.

The large improvement observed in this study may also reflect the nature of the task. Most ICD-coding studies focus on assigning multiple diagnoses from discharge summaries or electronic health records, which are often lengthy, complex, and highly variable. In contrast, our study focused on disease normalization of clinical trial conditions. These condition descriptions are generally shorter, more structured, and more focused on a specific disease. In addition, the LLM worked within a constrained decision framework supported by terminology mappings and semantic retrieval, which reduced the number of plausible codes and allowed the model to focus on selecting the most appropriate mention.

Overall, our findings should be interpreted within the broader context of recent advances in AI-assisted medical coding. Two conclusions emerge consistently from the literature. First, many apparent coding disagreements reflect clinical ambiguity rather than model failure. Second, the highest and most reliable performance is usually achieved when LLMs are combined with external knowledge sources, ontologies, knowledge graphs, or structured candidate-retrieval systems.

### Implications for disease-landscape of clinical trials analysis

The primary motivation for developing an automated disease-mapping pipeline is to perform a large-scale analysis of the disease landscape of German-affiliated clinical trials across multiple registries. Such analyses require the harmonization of condition descriptions from different trial registries, each of which uses its own terminology and coding system. Without a common classification framework, meaningful comparisons of research activity for different diseases across registries, trial designs, and time periods are difficult to achieve.

The results of this study demonstrate that automated harmonization of trial conditions to ICD-10 is feasible and can support population-level analyses of clinical trial activity. The deterministic pipeline achieved moderate-to-substantial reliability at the ICD-10 chapter and block levels, indicating that it can reliably identify broad disease groups and major therapeutic areas across registries.

The LLM-based evaluation provides evidence that more reliable three-character ICD-10 classification may be achievable. When fine-tuned and provided with clear and reliable context, the LLM could support future disease-landscape analyses in finer-grained detail than is currently possible with the deterministic pipeline alone.

### Limitations

The findings should be interpreted within the context of the respective trial registries and coding systems examined in this study. First, the pipeline was developed and evaluated using clinical trial registries and ICD-10 mappings relevant to this study. Its performance in other registries or healthcare datasets, or coding frameworks would require independent validation. Second, the LLM evaluation was performed using a single deterministic run (temperature = 0). As a result, the study did not assess variability in model outputs across repeated runs.

Third, the amount of trial information provided to the LLM was limited by predefined text-length restrictions. For a small number of trials with very long records, relevant contextual information may have been omitted, although the impact of this was not formally assessed. Fourth, recovery analyses for reject-band, review-band, and Tier-3 mentions were based on a stratified sample and not on the complete dataset.

Finally, the mention-extraction and splitting quality was not evaluated as a separate task; the gold-standard evaluation measures end-to-end accuracy against the expert reference, so errors originating specifically at the splitting stage cannot be distinguished from errors introduced during candidate generation or final code assignment.

## Conclusion

Among the methods evaluated, an LLM constrained to the same decision logic used to build the human reference standard achieved the highest agreement with expert coding (96.7% accuracy, κ = 0.966), substantially outperforming the deterministic ontology-and-embedding pipeline (49.0% accuracy, k = 0.487–0.490) at harmonizing health conditions across German-affiliated clinical trial registries onto a common ICD-10 classification. Future disease-landscape analyses of these registries could therefore use an LLM operating under the same constrained framework to map health conditions across these registries.

## Ethics statement

This study used only publicly available, aggregate clinical trial registration metadata; no individual patient data, human subjects, or animal research was involved. As the study did not involve human participants or personally identifiable information beyond publicly registered trial records, formal institutional review board (IRB) or ethics committee approval was not required.

## Conflicts of interest

Authors declare no conflicts of interest.

## Funding

No funding was acquired

## Author contributions

Conceptualization: RN, ALS. Methodology: RN, ALS. Software: RN. Validation: RN, EK. Formal analysis: RN. Data curation: RN, AF, DF, SGS, YSS. Writing and original draft: RN. Writing, review and editing: all authors.

## Data availability statement

This study used publicly available clinical trial registry data from the German Clinical Trials Register (DRKS), ClinicalTrials.gov, the EU Clinical Trials Register (EUCTR), and the Clinical Trials Information System (CTIS). All registry records used in this analysis are freely accessible through these platforms. The pipeline source code, mapping tables, and abbreviation dictionary described in this manuscript will be made available at https://github.com/RodrigueNdab/icd10_pipeline

## Appendix

## Appendix A. Formal specification of the mapping algorithm

### A.1 Notation and problem definition

Notation: Let mi denote a single condition mention extracted from a registry record (for example, “type 2 diabetes mellitus”). Let Tj denote trial j, and Mj = {m1, …, mk} denote the set of all condition mentions extracted from trial j after splitting. Let C = {c1, …, cn} denote the complete set of WHO ICD-10 (2019) three-character categories (for example, E11, C50, F32). Let C(mi) ⊆ C denote the set of candidate ICD codes generated for mention mi by the ontology and embedding pathways.

Goal: The mapping function assigns each mention to exactly one outcome: f: mi → {c*, REVIEW, REJECT}, where c* is the selected ICD-10 three-character code, REVIEW indicates a borderline mention that will be exported for manual adjudication, and REJECT indicates that no assignment reached sufficient confidence and the mention is retained with an explicit reason for rejection.

### A.2 Stage 2: Elastic-net ICD classifier, formal specification

Purpose: Stage 2 classifies each mention as either ICD-mappable (eligible for ICD-10 code assignment) or Tier 3 (a non-ICD concept such as healthy volunteer, pharmacokinetic study, or basic research).

Model: The probability that a mention is ICD-eligible is modelled using logistic regression: P(ICD_eligible | mi) = σ(β0 + xi^T^β)

where:

σ(·) is the logistic (sigmoid) function: σ(z) = 1 / (1 + exp(–z)), which maps any real-valued input to the range (0, 1) and can be interpreted as a probability.

β0 is the intercept (bias) term, representing the baseline log-odds of a mention being ICD-mappable when all features are zero.

xi is the feature vector for mention mi, comprising: (a) sparse TF-IDF word unigram and character n-gram (n = 2 – 4) representations of the mention text and any available contextual text (title, brief summary); (b) mention length in characters; (c) binary registry indicators (one per registry: DRKS, ClinicalTrials.gov, CTIS, EUCTR); and (d) primary purpose category (diagnostic, treatment, basic research, other).

β is the coefficient vector (one weight per feature), estimated from the training data. Each coefficient represents the change in log-odds of ICD-mappability associated with a one-unit increase in the corresponding feature.

Regularization: The coefficient vector is estimated via elastic-net regularization to prevent overfitting and handle the high-dimensional feature space:

β̂ = argminβ [ℓ(β) + λ(α||β||1 + (1 – α)||β||2²)] where:

ℓ(β) is the negative log-likelihood (logistic loss), measuring how well the model fits the training data. Minimizing ℓ alone would overfit the high-dimensional TF-IDF features.

λ is the regularization strength, controlling the trade-off between fitting the data and keeping coefficients small. A larger λ forces more coefficients toward zero. The optimal value is selected by 5-fold cross-validation.

α is the mixing parameter (set to 0.5), controlling the balance between two types of regularization: L1 (lasso, ||β||1 = Σ|βj|) which drives irrelevant features to exactly zero (sparsity), and L2 (ridge, ||β||2² = Σβj²) which shrinks correlated features smoothly rather than selecting one arbitrarily. At α = 0.5, both penalties contribute equally.

||β||1 = Σj|βj| is the L1 norm (sum of absolute values), encouraging sparse models where many coefficients are exactly zero.

||β||2² = Σjβj² is the squared L2 norm (sum of squared values), encouraging small but non-zero coefficients. Decision rule: ICD_eligible = 1 if P ≥ τ, and 0 otherwise. The threshold τ is calibrated on a held-out test set to maximize F1 on the minority (Tier 3) class.

Override rule: If mi contains a term from a curated disease keyword set (for example, cancer, diabetes, stroke, depression, asthma, obesity), then ICD_eligible = 1 regardless of the classifier output. This deterministic override prevents false negatives on unambiguous disease names.

### A.3 Stage 3: Deterministic candidate generation, formal specification

Purpose: Stage 3 generates a set of candidate ICD-10 codes for each ICD-mappable mention using deterministic ontology pathways.

Candidate set: C(mi) = CICD-GM ∪ CUMLS ∪ Clexical where:

CICD-GM = mapGM→WHO(mi) denotes the set of ICD-10 WHO codes obtained by converting ICD-10-GM codes from DRKS records to WHO equivalents using the official BfArM crosswalk table. This applies only to DRKS trials that have structured ICD-10-GM codes.

CUMLS denotes the set of ICD-10 codes obtained by mapping the mention text through the UMLS Metathesaurus. The pathway is: mi → CUI (Concept Unique Identifier) → ICD-10 codes. The CUI link is established by matching the normalized mention text against UMLS string tables for MeSH and MedDRA vocabularies. Exact string matches produce higher confidence candidates than partial or indirect matches.

Clexical denotes the set of ICD-10 codes obtained by approximate string matching, applied only when CICD-GM and CUMLS are both empty. Candidates are generated by computing Jaro-Winkler similarity (simJW) between the normalized mention and all ICD-10 WHO labels, retaining only those with simJW(mi, ck) ≥ δ, where δ = 0.80 is the minimum similarity threshold.

The union (∪) operator means that candidates from all applicable pathways are pooled and deduplicated. If the same ICD-10 code appears via multiple pathways (for example, E11 from both DRKS direct and UMLS exact), the highest confidence score is retained.

**Ontology candidate pathways and confidence scores.**

| Pathway | Registry scope | Method | Confidence | Role |
| --- | --- | --- | --- | --- |
| DRKS direct | DRKS | ICD-10-GM → WHO crosswalk | 1.00 | Highest-precision anchor |
| UMLS CUI exact | All | Exact UMLS string match → CUI → ICD-10 | 0.90 | High precision for known terms |
| UMLS CUI indirect | All | Substring/token match → CUI → ICD-10 | 0.70 | Broader recall, partial matches |
| MeSH hierarchy exact | ClinicalTrials.gov | Structured MeSH term parsing | 0.60 | Chapter-level evidence |
| MeSH hierarchy indirect | ClinicalTrials.gov | Indirect MeSH hierarchy match | 0.45 | Weak chapter-level signal |
| CTIS therapeutic area (TA) | CTIS | TA → ICD chapter mapping | 0.60 | Chapter constraint for CTIS |
| DRKS range prior | DRKS | Broad ICD-10-GM range evidence | 0.35 | Weak prior, never exact code |
| String fallback | All | Jaro-Winkler $\geq 0.80$ | 0.40 | Safety net for unmatched |

### A.4 Stage 4: Hybrid scoring, formal specification

For each candidate c_k_ ∈ C(m_i_), a hybrid confidence score combines ontology and embedding evidence. Two formulas are used, depending on whether ontology support exists.

For candidates with ontology support:

S(m_i_, c_k_) = 0.50·I_onto + 0.30·sim_emb + 0.20·sim_lex + b_exact + b_hybrid + b_tier1 − p_freq − p_vague − p_nondis + b_TA

For embedding-only candidates (no ontology support):

S(m_i_, c_k_) = 0.35·sim_emb + 0.55·sim_lex + 0.10·I_exact

where I_onto ∈ [0, 1] is the Stage 3 ontology confidence (zero for embedding-only candidates); sim_emb ∈ [0, 1] is SapBERT cosine similarity; sim_lex ∈ [0, 1] is Jaro–Winkler lexical similarity; I_exact ∈ {0, 1} indicates an exact string match; b_exact = 0.15 (ontology-backed formula only); b_hybrid = 0.03 when both ontology and embedding evidence support the same candidate; b_tier1 = 0.05 for Tier 1 disease-chapter candidates; p_freq is proportional to a candidate code’s log-frequency of prior assignment across the corpus; p_vague = 0.15, applied to a fixed blocklist of known attractor codes (C80, R69, Z00, D48, T98); p_nondis = 0.15, applied when the mention text itself matches a non-disease pattern; and b_TA (CTIS mentions only) = 0.08 or −0.10 depending on whether the candidate’s chapter agrees with the trial’s declared therapeutic area. (Full applied explanation and rationale: Methods, Stage 4.) Assignment. The selected code is c* = argmax_{c_k_ ∈ C(m_i_)} S(m_i_, c_k_) the candidate achieving the highest hybrid score. For example, given the mention “adult-onset diabetes” and three candidate codes-E10 (Type 1 diabetes), E11 (Type 2 diabetes), and E14 (unspecified diabetes) - the pipeline assigns whichever candidate achieves the highest composite score across ontology, embedding, and lexical evidence; in this illustrative case, E11 is selected as the best-supported match.

### A.5 Confidence decision rule

Each mention is assigned to one of three confidence bands based on the score of the winning candidate:

Decision(mi) = HIGH if S ≥ τh; REVIEW if τl ≤ S < τh; REJECT if S < τl

where τh is the high-confidence threshold and τl is the minimum-plausibility threshold. These thresholds will be calibrated on the gold-standard sample.

### A.6 Trial-level aggregation and evaluation metrics

Multi-label aggregation: The ICD-10 profile for trial j is the union of all high-confidence and review-band assignments: Yj = ⋃m_i_ ∈ Mⱼ f(mi). Each trial can therefore receive multiple ICD-10 codes (multi-label).

Mention-level metrics:

Precision = TP / (TP + FP) : of all mentions the pipeline assigned a code, how many were correct?

Recall = TP / (TP + FN) : of all mentions that should have received a code, how many did the pipeline find?

F1 = 2 × Precision × Recall / (Precision + Recall) : the harmonic mean, balancing precision and recall.

Inter-rater agreement: k = (Po – Pe) / (1 – Pe), where Po is the observed agreement rate between the two expert reviewers, and Pe is the agreement expected by chance. κ = 1 indicates perfect agreement; κ = 0 indicates agreement no better than chance.

Trial-level set similarity: Jaccard(Tj) = |Ypred ∩ Ytrue| / |Ypred ∪ Ytrue|, where ∩ is the intersection (codes in both predicted and true sets) and ∪ is the union (codes in either set). Jaccard = 1 means perfect agreement; Jaccard = 0 means no overlap.

## Appendix B LLM prompt specification

### B.1 Shared system prompt

This appendix specifies the attempted full prompt to be used for LLM validation tracks. The shared system prompt defines the coding process and rules; track-specific task headers differ only in framing (assign from scratch vs audit a candidate code) and output schema (assigned code vs comparison decision).

#### Shared system prompt

‘’You are a clinical coder assigning ICD-10 codes to condition mentions extracted from clinical-trial registry records. Your goal is to capture the disease being studied in the trial, not the surface form of the extracted mention.

Follow this decision cascade in order. Stop at the first step that yields a defensible answer.

STEP 1 : Mention itself

If CONDITION_MENTION is a recognisable disease or condition, assign the 3-character ICD-10 code directly. Example: a mention reading “type 2 diabetes mellitus” -> E11.

STEP 2: Infer from trial context

If the mention is fragmentary, vague, or not a disease on its own, read the OFFICIAL_TITLE and then FULL_TEXT and code the condition the trial is studying.

Before assigning a code at this step, state in one short sentence what condition the trial is studying.

Example: a mention consisting only of a procedural noun in a trial whose title identifies a specific musculoskeletal disorder -> code the disorder.

STEP 3a : Broad ICD-10 mapped to GBD Level 3

If a precise 3-character code cannot be defended but the disease area is clear, assign the broadest 3-character ICD-10 code that maps to a single GBD Level 3 cause, and write the GBD Level 3 cause name in notes.

Example: a trial in an unspecified solid tumour where the primary site is not given -> a 3-character code in the C-chapter that maps to a single GBD Level 3 neoplasm cause.

STEP 3b : GBD Level 3 name only

If no ICD-10 code (even broad) is defensible but a GBD Level 3 cause still fits, leave icd10_code blank and record the GBD Level 3 cause name in notes. STEP 3c: GBD Level 2 fallback

If even GBD Level 3 is too narrow, leave icd10_code blank and record the GBD Level 2 cause name in notes.

STEP 4: NO_ICD

Only if steps 1 to 3 all fail. Set no_icd = true and write “full check of the context” in notes. PREVENTION / HEALTHY-POPULATION RULE

If the trial tests prevention of a disease or health condition in a healthy population, code the disease or condition being prevented, not the population. Examples of the principle:

A behavioural intervention in healthy workers aimed at preventing a specific mental health outcome -> code in the mental and behavioural disorders chapter (F00-F99).

An intervention in healthy adults aimed at preventing a specific sexual function disorder -> code in the F52 block (sexual dysfunction not caused by organic disorder or disease).

NO_ICD is reserved for trials with no target condition at all (e.g. a Phase 1 pharmacokinetic study in healthy adults with no disease named). DISEASE OVER SYMPTOM AND AETIOLOGY OVER MANIFESTATION

When both appear, code the disease, not the symptom or manifestation.

A symptom mention (e.g. pain, fatigue) in a trial whose title makes the underlying disease explicit -> code the underlying disease.

A systemic disease that produces an organ-specific manifestation -> code the systemic disease unless the trial is explicitly limited to the manifestation.

A postoperative symptom in a trial focused on a specific underlying surgical indication -> code the underlying indication.’’

CONFIDENCE

high = step 1, or step 2 with one obvious condition.

medium = step 2 with interpretation, or step 3a with a clear GBD Level 3 area.

low = step 3b or 3c (GBD-name-only fallback).

OUTPUT

Return only a single JSON object matching the schema given in the task header. No prose outside the JSON

### B.2. Track A task and output schema

Track A ( review and reject bands)

TASK: Apply the cascade to the mention below and return your decision.

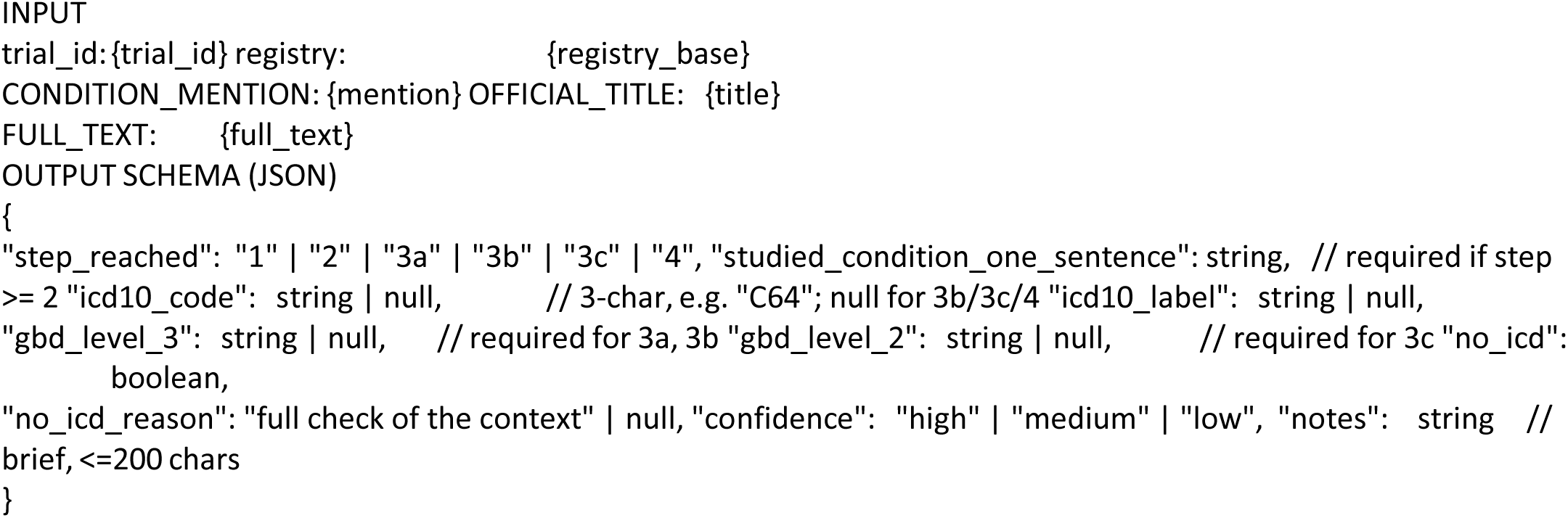

### B.3: Track B task and output schema

Track B (high-confidence band : audit pipeline candidate)

TASK: The pipeline has assigned a CANDIDATE ICD-10 code to the mention below. Apply the cascade INDEPENDENTLY, then compare your conclusion to the candidate.

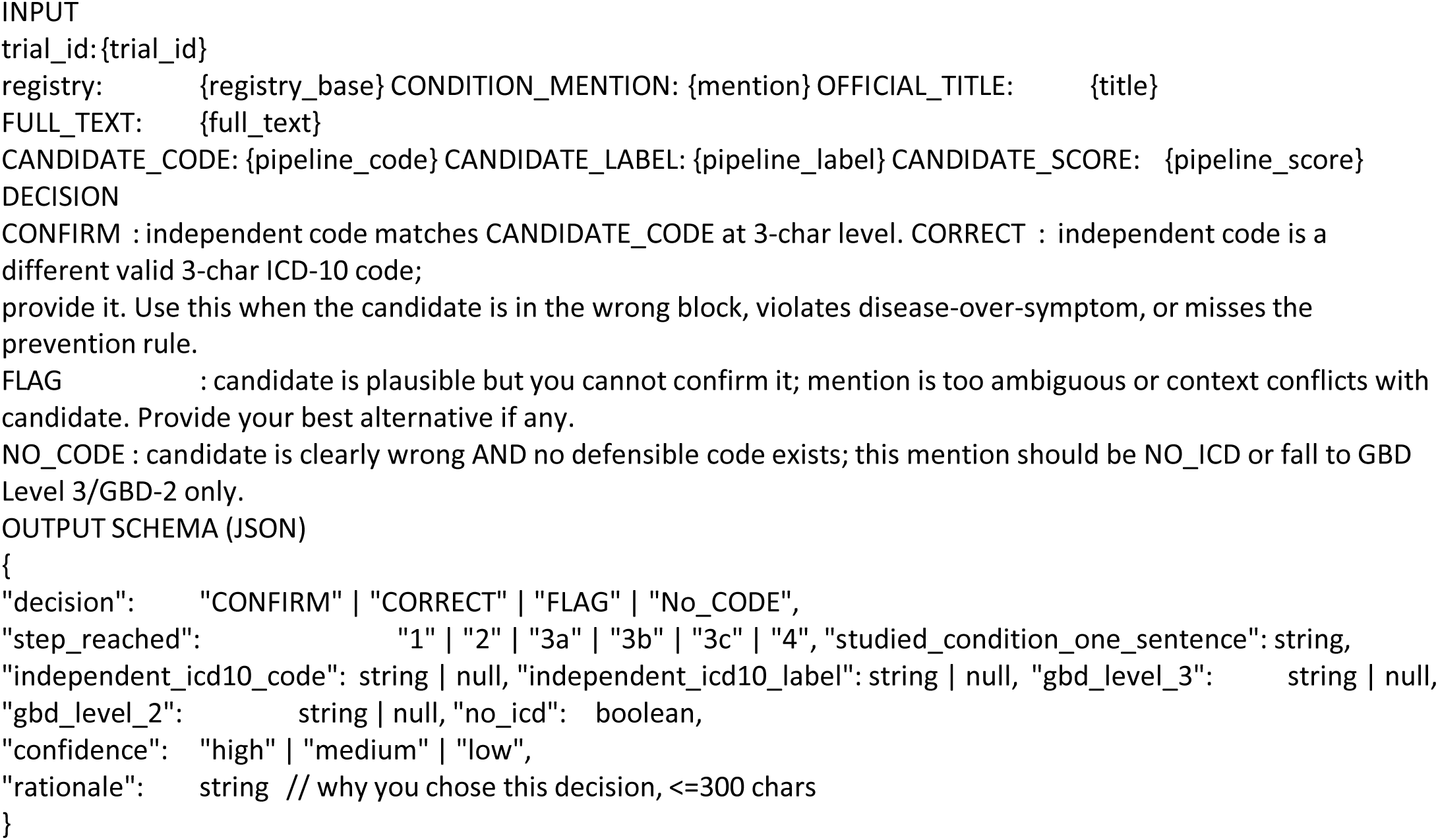

### B.4 Model and inference parameters

Model: gpt-4o (OpenAI); Temperature: 0; response_format: JSON.

### B.5 Human annotation guidance

This annotation guide mirrors the LLM system prompt (Appendix B.1) exactly, reflecting the shared decision cascade applied to both human reviewers and the LLM.

Goal: capture the disease being studied.

Step 1 : Mention itself If the CONDITION_MENTION is a recognisable disease/condition, assign the 3-char ICD-10 code directly. *Example:* “renal cell carcinoma” → C64.

Step 2 : Infer from trial context If the mention alone is insufficient or not clear (“transferring”, “any variant”, “appetite regulation”), read the official_title and then FULL_TEXT. Code the studied condition. *Example:* “transferring” in a HAL lumbar exoskeleton trial → M54 (dorsalgia).

Step 3a : Broad ICD-10 → GBD Level 3 If you cannot defend a precise 3-char code but the disease *area* is clear, give the broadest ICD-10 3-char code that maps to a single GBD Level 3 cause, and write the GBD Level 3 cause name in expert_notes. *Example:* unspecified solid tumour → C80 (“Other neoplasms”).

Step 3b : GBD Level 3 name only If no ICD-10 code (even broad) is defensible but a GBD Level 3 cause still fits, leave the ICD-10 code blank and record the GBD Level 3 cause name in expert_notes.

Step 3c : GBD Level 2 fallback If even GBD Level 3 is too narrow, record the GBD Level 2 cause name in expert_notes. Step 4 : NO_ICD only when none of the above hold. State in notes ‘full check of the context’ to indicate no health condition found after full check of the trial context

Prevention/healthy-population rule (important) If a trial tests prevention of a disease (or of a condition/health concept) in a healthy population, code the disease or condition being prevented, not the population.

“Stress prevention in staff nurses to prevent mental health problems” → code in the relevant mental health chapter/block (e.g. Mental and behavioural disorders (F00–F99)).

“Prevention of premature ejaculation in healthy males” → code the ejaculation condition (F52.4 → F52).

“Prevention of preterm birth in healthy pregnant women” → code O60 or Block: O60–O75 Complications of labour and delivery

Non_icd_10 is reserved for trials with no target condition at all (e.g. Phase 1 PK in healthy adults with no disease named)

Disease or symptom / aetiology over manifestation

Transthyretin amyloidosis with cardiomyopathy → E85, not I43.

Postoperative pain in a knee-replacement trial → M17 (the underlying disease being treated), not R52 or M25.

Confidence

high : Step 1, or Step 2 with one obvious condition.

medium : Step 2 with interpretation, or Step 3a with a clear GBD 3 area.

low : Step 3b/3c (GBD-name-only fallback)

## Notes

### Competing Interest Statement

The authors have declared no competing interest.

### Clinical Protocols

https://osf.io/mkn67/overview

### Author Declarations

The study used only publicly available clinical trial registry data. Sources included: ClinicalTrials.gov (U.S. National Library of Medicine): https://clinicaltrials.gov/ German Clinical Trials Register (DRKS): https://drks.de/search/en EU Clinical Trials Register (EUCTR): https://www.clinicaltrialsregister.eu/ctr-search/search Clinical Trials Information System (CTIS, European Union): https://euclinicaltrials.eu/ All data were publicly accessible clinical trial registry records. No registration, application, or special authorization was required to access the information.

