## Supplemental Data 1 for "Towards understanding the disease landscape of clinical trials in Germany: Ontology and embedding-based pipelines versus Large Language Models for ICD-10 Harmonization"

**Supplementary Note 1. Error analysis and mechanism**

**N1.1 False Positives and False Negatives.**

Six false positives in the SabBERT and four in method 2 were attributable to three primary mechanisms: (i) non diseases incorrectly interpreted as disease concepts, (ii) wellness-oriented language misclassified as pathological conditions, and (iii) embedding-attractor effects in COVID-related contexts, where semantic similarity to pandemic terminology led to incorrect mappings unrelated to the actual content of the mention. False negatives (n = 143) were identical across both methods because Stage 4 affects precision rather than coverage. These errors were concentrated almost exclusively in the embedding-only stratum (92/93) and the Tier-3-excluded stratum (51/51).

**N1.2 Ontology-Linking Failures.**

Ontology-linking failures were not recorded as a separate error category. Their frequency can be approximated by the ontology recall failure rate of 21.9% reported in the main text .

**N1.3 Embedding-Attractor errors.**

Embedding-attractor effects were illustrated by two COVID-context false positives, in which mentions relating to mask use and exercise metabolism were mapped to COVID-related codes on the basis of embedding similarity. These errors were identified qualitatively and were not systematically quantified across the full corpus.

**N1.4 Within-Family ambiguity.**

Within-family ambiguity constituted the largest residual error category, accounting for 20 of 56 errors in the baseline system and 23 of 56 in Method 2. Most cases involved confusion between a specific diagnostic subtype and an unspecified default code. Analysis suggests that these errors came from the pipeline's 0.03 tie-breaking threshold applied to competing candidates within the same ICD block.

**N1.5 Cross-chapter misclassification.**

Cross-chapter misclassifications accounted for 17 of 56 baseline errors and 16 of 56 Method 2 errors. These values are substantially lower than estimates derived from a naïve first-letter heuristic (26 and 22, respectively), which overestimates the phenomenon by treating any transition from C-coded to D-coded concepts as a chapter change, despite both ranges belonging to ICD-10 Chapter II.

**N1.6 Erroneous retention of non-condition text.**

Cases involving the incorrect retention of non-condition text overlapped directly with the false-positive mechanisms described in Section N1.1 and therefore do not constitute a distinct error category.

**N1.7 Candidate Recall.**

For the matched evaluation population, candidate recall was 78.1% (210/269) for the ontology pathway and 58.4% (157/269) for the embedding pathway. When both pathways were combined, recall reached 73.7% (249/338).

**Supplementary Table S1. Final outcome, full corpus, by support type.**

| **Support type** | **n** | **%** |
| --- | --- | --- |
| Ontology-supported | 31,896 | 80.7% |
| Hybrid (ontology + embedding agree) | 2,783 | 7.0% |
| Embedding-only | 4,451 | 11.3% |
| Whitelist rescue (curated override for known disease terms) | 382 | 1.0% |

**Supplementary Table S2. Final candidates by Stage 3 pathway.**

| **Pathway** | **n** | **%** |
| --- | --- | --- |
| DRKS direct (ICD-10-GM/WHO shared backbone) | 13,360 | 33.8% |
| MeSH (UMLS bridge, exact + indirect + hierarchy) | 16,224 | 41.1% |
| MedDRA (UMLS bridge, exact + indirect) | 3,285 | 8.3% |
| CTIS therapeutic-area | 851 | 2.2% |
| DRKS range-prior | 201 | 0.5% |
| String fallback | 71 | 0.2% |
| No ontology pathway (embedding-only) | 3,950 | 10.0% |

**Supplementary Table S3. Baseline vs. Method 2, corpus-wide confidence-band outcome.**

|  | **Baseline** | **Method 2** |
| --- | --- | --- |
| High-confidence | 28,604 (72.4%) | 28,368 (71.8%) |
| Review | 855 (2.2%) | 1,136 (2.9%) |
| Reject | 10,053 (25.4%) | 10,008 (25.3%) |

**Supplementary Table S4. Full pipeline confusion matrix (TP/FN/Wrong/FP/TN), baseline and method 2, all three hierarchy levels.**

| **Level** | **Method** | **TP** | **FN** | **Wrong** | **FP** | **TN** | **Accuracy** | **Precision** | **F1** | **κ** |
| --- | --- | --- | --- | --- | --- | --- | --- | --- | --- | --- |
| 3-character (n=390) | Baseline | 191 | 143 | 56 | 6 | 104 | 49.0% | 75.5% | 59.4% | 0.487 |
| 3-character (n=390) | Method 2 | 191 | 143 | 56 | 4 | 106 | 49.0% | 76.1% | 59.6% | 0.490 |
| Block (n=387) | Baseline | 211 | 143 | 33 | 6 | 104 | 54.5% | 84.4% | 66.2% | 0.533 |
| Block (n=387) | Method 2 | 214 | 143 | 30 | 4 | 106 | 55.3% | 86.3% | 67.4% | 0.543 |
| Chapter (n=387) | Baseline | 227 | 143 | 17 | 6 | 104 | 58.7% | 90.8% | 71.3% | 0.561 |
| Chapter (n=387) | Method 2 | 228 | 143 | 16 | 4 | 106 | 58.9% | 91.9% | 71.8% | 0.567 |

**Supplementary Table S5. Error taxonomy, hierarchy-corrected.**

| **Category** | **Baseline (naïve → corrected)** | **Method 2 (naïve → corrected)** |
| --- | --- | --- |
| Within-family | 15 → 20 | 17 → 23 |
| Within-chapter | 15 → 16 | 17 → 14 |
| Cross-chapter | 26 → 17 | 22 → 16 |

**Supplementary Table S6. Baseline accuracy by design stratum.**

| **Design stratum** | **n** | **Baseline accuracy** |
| --- | --- | --- |
| Ontology-supported | 197 | 79.2% |
| Unspecified default | 49 | 69.4% |
| Embedding-only | 93 | 1.1% |
| Tier-3 excluded | 51 | 0.0% (structural) |
